# No Evidence for Genetic Role of the Tumor Necrosis Factor Pathway in Parkinson’s Disease

**DOI:** 10.1101/2025.09.06.25335210

**Authors:** Morvarid Ghamgosar Shahkhali, Lang Liu, Emma N. Somerville, Alastair J Noyce, Ziv Gan-Or, Konstantin Senkevich

## Abstract

Tumor necrosis factor (TNF) inhibition is under investigation as a therapeutic strategy for Parkinson’s disease (PD) and REM sleep behavior disorder (RBD), yet supporting genetic evidence is limited. We used Summary-data-based Mendelian Randomization (SMR) to test whether expression level of ten TNF-related genes were causally linked to PD risk, PD progression, or RBD risk. We also analyzed associations between common and rare variants in these genes, and performed pathway specific polygenic risk score analysis, with PD. Overall, our findings do not support a genetic link between the TNF signaling and PD or RBD, arguing against this pathway as a genetically validated therapeutic target.

## Introduction

Neuroinflammation contributes to dopaminergic neuron loss and the progression of Parkinson’s disease (PD)^1,2^. Neuronal degeneration in the substantia nigra triggers microglial activation, driving pro-inflammatory cytokine release^3^. Among these mediators, tumor necrosis factor (TNF) plays a central role^4^. Sustained inflammatory signaling perpetuate itself, creating a vicious cycle that may accelerate PD progression^5^.

TNF and its downstream signaling cascade have been consistently implicated in neuroinflammatory responses and dopaminergic neurodegeneration in cellular and animal models of PD^4,6,7^. However, the genetic evidence linking TNF-related genes to PD risk remains limited and inconclusive^7–10^. To date, no genes within the TNF pathway have been identified as genome-wide significant loci (p ≤ 5 × 10⁻D) in PD or REM sleep behavior disorder (RBD), a prodromal clinical marker of PD and other synucleinopathies^11–13^. Moreover, recent Mendelian randomization (MR) studies have found no evidence supporting a causal role of TNF signaling in modifying PD risk or age at onset^14,15^. Nevertheless, there is ongoing clinical interest, and early-phase clinical trials targeting TNF signaling in PD and RBD are underway (https://clinicaltrials.gov/study/NCT06996652)^16^, underscoring the need for robust human genetic evidence to support or refute this therapeutic strategy.

In this study, we investigated the genetic contribution of key components of the TNF signaling cascade to PD and RBD. Specifically, we used Summary-data-based MR (SMR) to test whether genetically driven expression of the key TNF-pathway genes (*TNF*, *TNFRSF1A*, *TNFRSF1B*, *TRADD*, *TRAF1*, *TRAF2*, *TRAF5*, *CASP8*, *NFKB1*, and *NFKB2*; Supplementary Table 1) affects PD risk and progression, and RBD risk. In parallel, we evaluated associations of common and rare variants in these genes, together with pathway-level analyses, for association with PD risk.

## Results

### No evidence for causal associations between TNF-pathway gene expression and disease

To explore potential causal relationships between gene expression and disease risk or progression, we performed SMR using expression quantitative trait loci (eQTL) data from caudate, cortex, substantia nigra, and whole blood. *TRAF5* eQTLs in brain cortex showed a nominal association with PD risk (P = 0.048, Pfdr = 0.607), while *CASP8* showed nominal associations with RBD risk in both brain cortex and whole blood (P = 0.033, Pfdr = 0.607; P = 0.002, Pfdr = 0.102, respectively). None of the associations of TNF-pathway genes remained significant (P < 0.05) after false discovery rate (FDR) correction (Supplementary Table 2). eQTL data for *TNF*, *TNFRSF1B*, and *TRADD,* were unavailable in studied tissues and therefore could not be tested. Overall, the SMR results do not support a causal role between TNF-related gene expression in PD or RBD.

### Rare and common variant analyses of TNF-related genes revealed no significant associations

We investigated the association between rare variants in TNF-related genes and PD using optimized Sequence Kernel Association Test (SKAT-O) across the Accelerating Medicines Partnership – Parkinson’s Disease (AMP-PD), UK Biobank (UKB) including UKB-PD (PD cases and controls), and UKB-PROXY (PD cases, proxy-cases and controls) cohorts. Meta-analyses of AMP-PD and UKB-PD revealed nominal significance for *TRAF5* variants with high Combined Annotation Dependent Depletion (CADD) scores and missense variants (P = 0.005, Pfdr = 0.312; and P = 0.037, Pfdr = 0.598, respectively), which were consistent with results from the AMP-PD and UKB-PROXY meta-analysis for high-CADD variants (P = 0.032, Pfdr = 0.418). No gene-level associations remained significant (P < 0.05) after multiple testing correction. At the pathway level, combining all studied genes together, no associations were observed across any functional variant groups (Supplementary Tables 3 and 4).

LocusZoom visualization of common variants (minor allele frequency (MAF) >D0.01) in TNF-related genes using the multi-ancestry PD Genome-Wide Association Study (GWAS) summary statistics revealed no significant associations (p ≤ 5 × 10⁻D) with PD risk (Figure 1).

**Figure 1.**
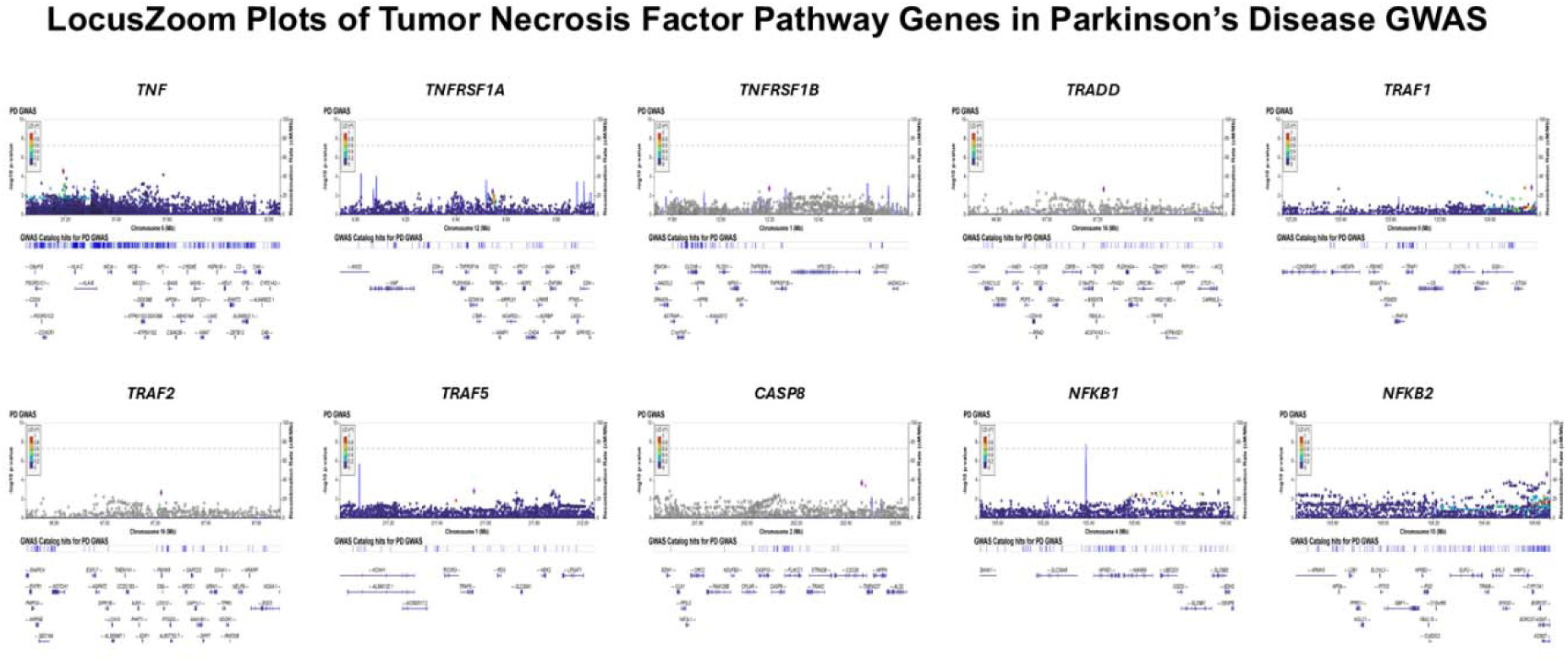
LocusZoom plots of *TNF, TNFRSF1A, TNFRSF1B, TRADD, TRAF1, TRAF2, TRAF5, CASP8, NFKB1,* and *NFKB2* genes in Parkinson’s disease genome-wide association study (GWAS). The lead SNP (i.e., the variant with the lowest P-value within each gene) is shown as a purple square. Variants are colored based on their linkage disequilibrium (LD) with the lead SNP, while grey variants indicate SNPs with no available LD information. The dashed horizontal line marks the genome-wide significance threshold (P < 5 × 10⁻D). X-axis: chromosomal position (Mb); Y-axis: –log₁₀(P-value) from GWAS.

### No evidence for polygenic effect of TNF-pathway variants on PD risk

We conducted pathway-based Polygenic risk score (PRS) analysis for the ten TNF-related genes using data from 15,663 individuals with PD and 76,551 controls across seven cohorts (Supplementary Table 5). In the McGill cohort, higher PRS was significantly associated with reduced risk of PD risk (OR = 0.895, 95% CI = 0.841–0.953, P = 4.9×10⁻D). However, this effect was not replicated in other cohorts or in the meta-analysis (Figure 2; Supplementary Table 6), and is therefore likely spurious.

**Figure 2.**
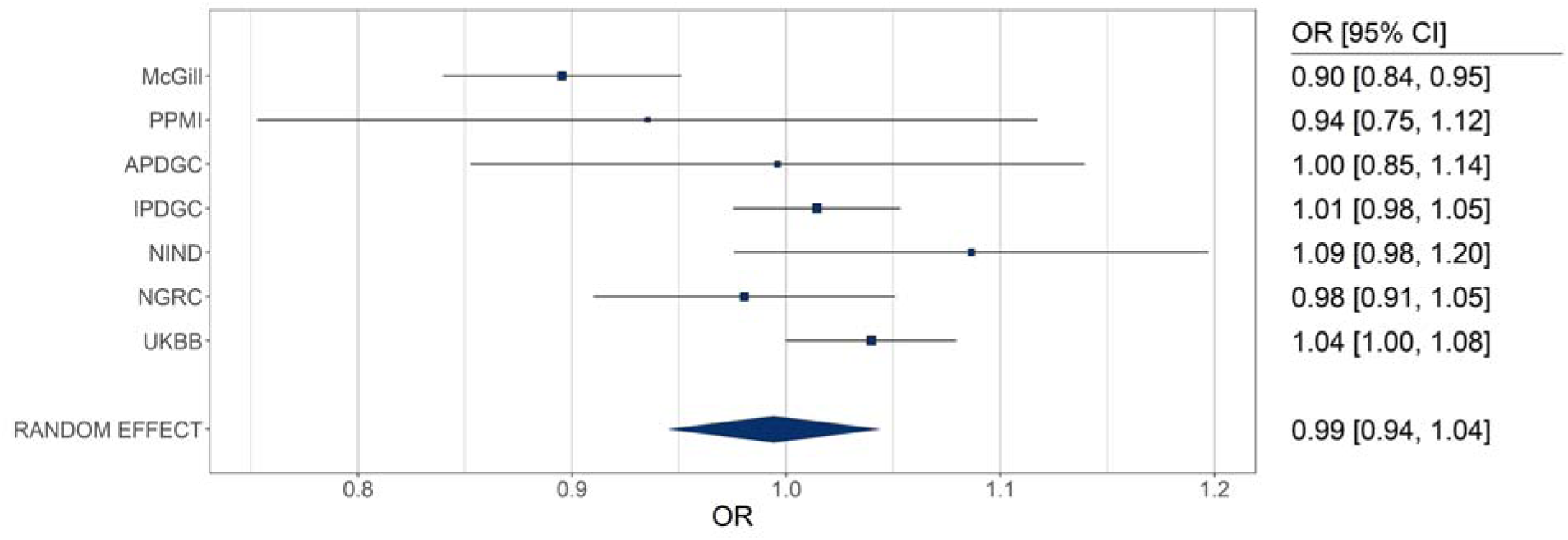
Pathway specific polygenic risk score analysis of TNF genes. OR-odds ratio; CI- confidence interval; PPMI- Parkinson’s Progression Markers Initiative; APDGC- Autopsy-Confirmed Parkinson Disease GWAS Consortium; IPDGC- International Parkinson’s Disease Genomics Consortium; NINDS- National Institute of Neurological Disorders and Stroke Genome-wide genotyping in Parkinson’s disease, NGRC- NeuroGenetics Research Consortium; UKBB- UK Biobank.

## Discussion

We found no evidence of a causal relationship between the expression of TNF signaling pathway genes and PD, including risk and progression or RBD risk. Additionally, rare and common variant analyses, and TNF-pathway PRS revealed no significant associations with PD risk.

The role of TNF signaling in PD remains controversial. Several studies have reported elevated serum TNF levels in PD and isolated RBD patients compared to controls^17–20^, suggesting a potential pro-inflammatory role for TNF in disease pathogenesis. However, other studies failed to consistently replicate these findings^21,22^. Although, higher serum levels of soluble TNFR1 were reported in PD patients relative to controls^21^. Importantly, it remains unclear whether elevated TNF reflects a causal factor contributing to disease onset and progression, or instead represents a downstream consequence of neurodegeneration and cell death. A cohort study of individuals with inflammatory bowel disease indicated that early anti-TNF therapy may reduce PD risk, supporting a possible protective role^23^. In parkinsonism animal models, blocking TNF signaling in the substantia nigra has been shown to attenuate dopaminergic neuron loss and associated behavioral deficits^24,25^. Yet, in two recently published α-synuclein-driven parkinsonism models, soluble TNF inhibition did not reduce α-synuclein pathology, neuroinflammation, or dopaminergic neurodegeneration ^26,27^.

Despite limited genetic evidence, TNF inhibition is currently being investigated in clinical trials as a potential therapeutic approach for PD and RBD (https://clinicaltrials.gov/study/NCT06996652)^16^. A MR study found no evidence that inhibition of TNF–TNFR1 signaling reduces PD risk or delays disease onset^14^, consistent with our own genetic findings. We assessed the association of TNF pathway with PD and RBD more comprehensively by including ten key pathway genes and evaluating both individual gene associations and the combined effect of the pathway.

Our study has several limitations. First, the sample sizes of some datasets, particularly for eQTLs from brain tissues, may have limited statistical power, especially for genes with low or tissue-specific expression. In addition, eQTL data for three genes were unavailable in tissues studied and therefore could not be tested with SMR. Another limitation is that brain mRNA– protein correlations are modest and protein QTL (pQTLs) often do not overlap eQTLs^28^. As a result, eQTL-based analyses may miss protein-level effects in TNF-pathway genes. Although we used two large independent biobank datasets for rare variant analysis, the study may still be underpowered to detect modest effect sizes. Our reliance on genetic and transcriptomic data alone does not capture post-transcriptional regulation or protein-level effects, or the influence of clinical factors such as medication use (e.g., levodopa or dopaminergic agonists), which may modulate TNF pathway activity in PD. Future studies incorporating more deeply phenotyped cohorts and proteomic integration will be essential to fully assess the role of immune signaling in PD.

In conclusion, we found no genetic evidence supporting a role for TNF signaling in PD or RBD, suggesting it may not represent a genetically supported therapeutic target. These results underscore the importance of integrating genetic data into therapeutic development, as studies lacking such guidance are more likely to fail^29^.

## Methods

### Study populations

For the SMR analysis^30^, we used summary-level eQTL data for ten TNF-related genes (Supplementary Table 1). These genes are the core mediators of TNF receptor signaling and regulate inflammation, cell survival, and cell death. Exposures were BrainMeta v.2^31^ cortex cis-eQTL (N=2,443), and Genotype-Tissue Expression (GTEx) v.8^32^ cis-eQTL across caudate (N individuals = 194), brain cortex (N individuals = 205), substantia nigra (N individuals = 114), and whole blood (N individuals = 670). Outcomes included GWAS summary statistics of PD motor and cognitive progression, Montreal Cognitive Assessment (MoCA) and the Unified Parkinson’s Disease Rating Scale (UPDRS) Part III (N cases = 4,093)^33^, and a PD motor progression (N cases = 6,766) ^34^ for which genome-wide survival analyses were conducted using time to motor progression endpoint, defined by reaching Hoehn and Yahr stage 3 or greater. In addition, GWAS summary statistics of PD risk (N cases = 63,555; N proxy-cases = 17,700; N controls = 1,746,386)^12^ and isolated RBD risk (N cases = 1,061; N controls = 8,386)^13^ were included as outcomes. Linkage disequilibrium (LD) score reference data from the 1000 Genomes Project Phase 3 for European ancestry (https://zenodo.org/records/10515792) were used in the analysis to account for LD patterns among single-nucleotide polymorphism (SNPs).

To analyze rare variants, we used whole genome sequencing data of unrelated individuals of European ancestry from two dataset: the UKB^35^ and the AMP-PD initiative cohorts (https://amp-pd.org/; detailed in the Acknowledgment). In the UKB, two cohorts were defined. The first cohort, hereafter referred to as the UKB-PD cohort, consisted of 3,134 PD cases and 60,171 randomly selected controls, after excluding individuals with diagnoses of mental and behavioral disorders (codes F00–F99) or nervous system diseases (codes G00–G99), based on International Classification of Diseases version-10 (ICD-10, field 41270). The second cohort, hereafter referred to as the UKB-PROXY cohort, included the same samples as the first, with the addition of PD proxy cases (healthy individuals with a first-degree relative with PD), to enhance the power to detect associations (N cases = 17,170, and N controls = 60,171). The AMP-PD cohort included 1,931 PD cases and 3,062 controls.

For common variant visualization, we used multi-ancestry PD GWAS (N cases = 49,049; N proxy cases = 18,785; N controls = 2,458,063)^36^.

For PRS pathway analysis, we used the genotyping data from 7 cohorts (detailed in Supplementary Table 5): McGill, Parkinson’s Progression Markers Initiative (PPMI), Autopsy-Confirmed Parkinson Disease GWAS Consortium (APDGC) (dbGaP phs000394.v1.p1), International Parkinson’s Disease Genomics Consortium (IPDGC) NeuroX dataset (dbGap phs000918.v1.p1), National Institute of Neurological Disorders and Stroke (NINDS) Genome-wide genotyping in Parkinson’s disease (dbGap phs000089.v4.p2), NeuroGenetics Research Consortium (NGRC) (dbGap phs000196.v3.p1) and UKB. Effect size estimates were derived from the most recent largest European PD GWAS^12^.

### Summary-data-based Mendelian randomization

We applied SMR analysis to evaluate whether expression of the ten TNF-pathway genes have a causal effect on PD and RBD traits. Significant cis-eQTLs (p ≤ 5 × 10⁻D) from BrainMeta and GTEx datasets were selected based on their location within or near each gene, using cis-window thresholds of ±500 kb. The HEIDI test was used to distinguish true causal effects from those due to linkage. To control for multiple testing, FDR correction^37^ was applied to reduce the likelihood of false-positive findings.

### Rare variant association analysis

We performed rare variant association analysis for TNF-related genes to investigate their potential contribution to PD risk. From the ten genes, we first selected rare variants (MAF ≤D0.01) in all cohorts. For the UKB data specifically, variants were further restricted to biallelic sites with sufficient depth of coverage (DPD≥D25) and low genotype missingness (missingness <D0.05), to match the quality control criteria used in the AMP-PD dataset (https://amp-pd.org/data/genomic-data). Variant annotation was performed using the Ensembl Variant Effect Predictor (Ensembl VEP, v.107.0)^38^ with the plugins noted at (https://github.com/saeidamiri1/bioinformatics_material/blob/main/docs/drac/posts/vep.md).

Variants were categorized into three groups: (1) missense variants, (2) loss-of-function (LoF) variants, including stop gained, frameshift, start lost, stop lost, splice acceptor, splice donor, transcript ablation, and exon loss variants, and (3) highly predicted deleterious variants, defined as variants with CADD scores greater than 20. For each cohort, SKAT-O was performed to test rare variants and functional variants in each gene using SKATBinary function^39^ in SKAT^40^. Moreover, at the pathway level, we evaluated the combined effect of all genes within each functional group. To combine results across AMP-PD cohort and each of the UKB cohorts, we performed meta-analysis using MetaSKAT^41^. All association tests were adjusted for age, sex, and the first five principal components. P-values were corrected for multiple testing using the FDR method^37^.

### Common variant association visualization

We generated LocusZoom plots (http://locuszoom.org/)^42^ for common variants (MAF >D0.01) at TNF-pathway loci using the multi-ancestry PD GWAS^36^.

### Pathway polygenic risk score analysis

To investigate the potential genetic association of the TNF pathway with PD, we calculated pathway PRS using PRSet^43^ for the TNF-related genes, with effect size estimates from most recent PD GWAS^12^. The analysis was restricted to individuals of European ancestry, excluding first- and second-degree relatives. Sex discrepancy analysis was performed by comparing recorded biological sex with genetically inferred sex, based on X chromosome heterozygosity and homozygosity rates, using the --check-sex function in PLINK v.1.9. Samples with discordant sex information—defined as X chromosome homozygosity estimates >0.85 for males and <0.25 for females—were excluded to improve data quality. Only common SNPs (MAF > 0.01) were retained. LD clumping was applied to remove variants with r²D>D0.1 within a 250 kb window. Covariates included age at onset (for cases), age at enrollment (for controls), sex, and the top five principal components.

## Data Availability

eQTL data from BrainMeta version 2 cis-eQTL and Genotype-Tissue Expression (GTEx) version 8 cis-eQTL summary statistics are available at https://yanglab.westlake.edu.cn/software/smr/#DataResource. GWAS summary statistics for PD progression, the Montreal Cognitive Assessment (MoCA), and UPDRS Part 3 traits are publicly available at https://pdgenetics.shinyapps.io/pdprogmetagwasbrowser/. Summary statistics for the PD motor progression trait can be downloaded from https://tinyurl.com/PDprogressionv2 (DOI: 10.5281/zenodo.8017385). Full PD risk GWAS summary statistics (GP2 2025) are publicly available at https://ndkp.hugeamp.org/research.html?pageid=a2f_downloads_280. Multi-ancestry PD risk GWAS summary statistics (Kim et al. 2024) are accessible via the Neurodegenerative Disease Knowledge Portal at https://ndkp.hugeamp.org/. The iRBD summary statistics can be found in the GWAS Catalog (https://www.ebi.ac.uk/gwas/, study accession GCST90204200). The data used in the preparation of this article were obtained from the AMP PD Knowledge Platform (https://www.amp-pd.org) and the UKBB via Neurohub (https://www.mcgill.ca/hbhl/neurohub).

## Code Availability

The code supporting the findings of this study will be made publicly available on GitHub upon publication at: https://github.com/mghamg/TNF-Study.

## Data Availability

eQTL data from BrainMeta version 2 cis-eQTL and Genotype-Tissue Expression (GTEx) version 8 cis-eQTL summary statistics are available at https://yanglab.westlake.edu.cn/software/smr/#DataResource. GWAS summary statistics for PD progression, the Montreal Cognitive Assessment (MoCA), and UPDRS Part 3 traits are publicly available at https://pdgenetics.shinyapps.io/pdprogmetagwasbrowser/. Summary statistics for the PD motor progression trait can be downloaded from https://tinyurl.com/PDprogressionv2 (DOI: 10.5281/zenodo.8017385). Full PD risk GWAS summary statistics (GP2 2025) are publicly available at https://ndkp.hugeamp.org/research.html?pageid=a2f_downloads_280. Multi-ancestry PD risk GWAS summary statistics (Kim et al. 2024) are accessible via the Neurodegenerative Disease Knowledge Portal at https://ndkp.hugeamp.org/. The iRBD summary statistics can be found in the GWAS Catalog (https://www.ebi.ac.uk/gwas/, study accession GCST90204200). The data used in the preparation of this article were obtained from the AMP PD Knowledge Platform (https://www.amp-pd.org) and the UKBB via NeuroHub (https://www.mcgill.ca/hbhl/neurohub).

https://pdgenetics.shinyapps.io/pdprogmetagwasbrowser/

https://tinyurl.com/PDprogressionv2

https://ndkp.hugeamp.org/research.html?pageid=a2f_downloads_280

https://ndkp.hugeamp.org/

https://www.ebi.ac.uk/gwas/

https://www.mcgill.ca/hbhl/neurohub

## Acknowledgments

We would like to thank the research participants for contributing to this study. MGS is supported by a graduate student award, Jeanne Timmins Costello Award. ZGO is supported by the Fonds de recherche du Québec–Santé (FRQS) Chercheurs-boursiers award and is a William Dawson Scholar. This study was supported by The Canadian Consortium on Neurodegeneration in Aging (CCNA), and by the GBA1-Canada initiative (G-Can - https://gba1can.org/), which is funded by The Hilary and Galen Weston Foundation and The Van Berkom family. Data used in the preparation of this article were obtained from the Accelerating Medicine Partnership® (AMP®) Parkinson’s Disease (AMP PD) Knowledge Platform. For up-to-date information on the study, visit https://www.amp-pd.org. The AMP® PD program is a public-private partnership managed by the Foundation for the National Institutes of Health and funded by the National Institute of Neurological Disorders and Stroke (NINDS) in partnership with the Aligning Science Across Parkinson’s (ASAP) initiative; Celgene Corporation, a subsidiary of Bristol-Myers Squibb Company; GlaxoSmithKline plc (GSK); The Michael J. Fox Foundation for Parkinson’s Research; Pfizer Inc.; AbbVie Inc.; Sanofi US Services Inc.; and Verily Life Sciences. Accelerating medicines partnership and AMP are registered service marks of the U.S. Department of Health and Human Services. Clinical data and biosamples used in preparation of this article were obtained from the (i) Michael J. Fox Foundation for Parkinson’s Research (MJFF) and National Institutes of Neurological Disorders and Stroke (NINDS) BioFIND study, (ii) Harvard Biomarkers Study (HBS) and the Stephen & Denise Adams Center for Parkinson’s Disease Research of Yale School of Medicine (CPDR-Y), (iii) National Institute on Aging (NIA) International Lewy Body Dementia Genetics Consortium Genome Sequencing in Lewy Body Dementia Case-control Cohort (LBD), (iv) MJFF LRRK2 Cohort Consortium (LCC), (v) NINDS Parkinson’s Disease Biomarkers Program (PDBP), (vi) MJFF Parkinson’s Progression Markers Initiative (PPMI), and (vii) NINDS Study of Isradipine as a Disease-modifying Agent in Subjects With Early Parkinson Disease, Phase 3 (STEADY-PD3) and (viii) the NINDS Study of Urate Elevation in Parkinson’s Disease, Phase 3 (SURE-PD3). BioFIND is sponsored by The Michael J. Fox Foundation for Parkinson’s Research (MJFF) with support from the National Institute for Neurological Disorders and Stroke (NINDS). The BioFIND Investigators have not participated in reviewing the data analysis or content of the manuscript. For up-to-date information on the study, visit michaeljfox.org/news/biofind. Genome sequence data for the Lewy body dementia case-control cohort were generated at the Intramural Research Program of the U.S. National Institutes of Health. The study was supported in part by the National Institute on Aging (program #: 1ZIAAG000935) and the National Institute of Neurological Disorders and Stroke (program #: 1ZIANS003154). The Harvard Biomarker Study (HBS) is a collaboration of HBS investigators [full list of HBS investigators found at https://www.bwhparkinsoncenter.org/biobank/ and funded through philanthropy and NIH and Non-NIH funding sources. The Stephen & Denise Adams Center for Parkinson’s Disease Research of Yale School of Medicine is funded through philanthropy and NIH and non-NIH funding sources. The HBS and CPDR-Y Investigators have not participated in reviewing the data analysis or content of the manuscript. Data used in preparation of this article were obtained from The Michael J. Fox Foundation sponsored LRRK2 Cohort Consortium (LCC). The LCC Investigators have not participated in reviewing the data analysis or content of the manuscript. For up-to-date information on the study, visit https://www.michaeljfox.org/biospecimens). PPMI is sponsored by The Michael J. Fox Foundation for Parkinson’s Research and supported by a consortium of scientific partners: [list the full names of all of the PPMI funding partners found at https://www.ppmi-info.org/about-ppmi/who-we-are/study-sponsors]. The PPMI investigators have not participated in reviewing the data analysis or content of the manuscript. For up-to-date information on the study, visit www.ppmi-info.org. The Parkinson’s Disease Biomarker Program (PDBP) consortium is supported by the National Institute of Neurological Disorders and Stroke (NINDS) at the National Institutes of Health. A full list of PDBP investigators can be found at https://pdbp.ninds.nih.gov/policy. The PDBP investigators have not participated in reviewing the data analysis or content of the manuscript. The Study of Isradipine as a Disease-modifying Agent in Subjects With Early Parkinson Disease, Phase 3 (STEADY-PD3) is funded by the National Institute of Neurological Disorders and Stroke (NINDS) at the National Institutes of Health with support from The Michael J. Fox Foundation and the Parkinson Study Group. For additional study information, visit https://clinicaltrials.gov/ct2/show/study/NCT02168842. The STEADY-PD3 investigators have not participated in reviewing the data analysis or content of the manuscript. The Study of Urate Elevation in Parkinson’s Disease, Phase 3 (SURE-PD3) is funded by the National Institute of Neurological Disorders and Stroke (NINDS) at the National Institutes of Health with support from The Michael J. Fox Foundation and the Parkinson Study Group. For additional study information, visit https://clinicaltrials.gov/ct2/show/NCT02642393.

The SURE-PD3 investigators have not participated in reviewing the data analysis or content of the manuscript. This research used the NeuroHub infrastructure and was undertaken thanks in part to funding from the Canada First Research Excellence Fund, awarded through the Healthy Brains, Healthy Lives initiative at McGill University, Calcul Québec and Compute Canada. This research has been conducted using the UK Biobank Resource under Application Number 45551. The UKBB cohort was accessed using Neurohub (https://www.mcgill.ca/hbhl/neurohub).

## Author Contributions

MGS conducted the analysis, interpreted the results, and wrote the manuscript. LL and ENS assisted with computational analysis and support. AJN and KS conceived the research idea and developed the methodology. ZG-O and KS supervised the study and contributed to the interpretation of the results. All authors reviewed, approved and contributed to editing the final manuscript.

## Competing Interests

ZG-O received consultancy fees from Lysosomal Therapeutics Inc. (LTI), Idorsia, Prevail Therapeutics, Ono Therapeutics, Denali, Handl Therapeutics, Neuron23, Bial Biotech, Bial, UCB, Capsida, Vanqua bio, Congruence Therapeutics, Takeda, Jazz Guidepoint, Lighthouse and Deerfield. The remaining authors declare no competing interests. AJN reports grants from Parkinson’s UK, Barts Charity, Cure Parkinson’s, National Institute for Health and Care Research, Innovate UK, the Medical College of Saint Bartholomew’s Hospital Trust, Alchemab, Aligning Science Across Parkinson’s Global Parkinson’s Genetics Program (ASAP-GP2) and the Michael J Fox Foundation. AJN reports fees from AstraZeneca, AbbVie, Bial, and Britannia. KS reports grants from Parkinson Canada, Aligning Science Across Parkinson’s Global Parkinson’s Genetics Program (ASAP-GP2) and is a consultant for Acurex.

## Notes

### Competing Interest Statement

Z.G-O. received consultancy fees from Lysosomal Therapeutics Inc. (LTI), Idorsia, Prevail Therapeutics, Ono Therapeutics, Denali, Handl Therapeutics, Neuron23, Bial Biotech, Bial, UCB, Capsida, Vanqua bio, Congruence Therapeutics, Takeda, Jazz Guidepoint, Lighthouse and Deerfield. The remaining authors declare no competing interests. A.J.N. reports grants from Parkinson's UK, Barts Charity, Cure Parkinson's, National Institute for Health and Care Research, Innovate UK, the Medical College of Saint Bartholomew's Hospital Trust, Alchemab, Aligning Science Across Parkinson's Global Parkinson's Genetics Program (ASAP-GP2) and the Michael J Fox Foundation. A.J.N. reports fees from AstraZeneca, AbbVie, Bial, and Britannia. K.S. reports grant from Parkinson Canada, Aligning Science Across Parkinson's Global Parkinson's Genetics Program (ASAP-GP2) and is a consultant for Acurex.

### Author Declarations

Ethics approval for the research study was granted by the McGill University Research Ethics Board.

