## Supplementary Material for "No Evidence for Genetic Role of the Tumor Necrosis Factor Pathway in Parkinson’s Disease"

Supplementary Table 1. Genes in the TNF signaling pathway and their functions

| Gene | Full Gene Name | Function |
|---|---|---|
| *TNF* | Tumor Necrosis Factor | Encodes TNF-α, a pro-inflammatory cytokine mediating neuroinflammation. |
| *TNFRSF1A* | TNF Receptor Superfamily Member 1A | Encodes TNFRSF1A (TNFR1), the main receptor for TNF; triggers apoptosis or NF-κB-mediated survival/inflammatory signaling. |
| *TNFRSF1B* | TNF Receptor Superfamily Member 1B | Encodes TNFRSF1B (TNFR2), a TNF receptor mainly promoting cell survival and immune regulation via NF-κB activation. |
| *TRADD* | TNFRSF1A-associated via death domain | Encodes TRADD, an adaptor protein recruited by TNFR1; involved in NF-κB activation or, alternatively, apoptosis. |
| *TRAF2* | TNF Receptor Associated Factor 2 | Encodes TRAF2, an E3 ubiquitin ligase recruited by TNFR1/TRADD, contributes to NF-κB activation. |
| *TRAF5* | TNF Receptor Associated Factor 5 | Encodes TRAF5, contributes to NF-κB signaling. |
| *TRAF1* | TNF Receptor Associated Factor 1 | Encodes TRAF1, which together with TRAF2 contributes to NF-κB activation. |
| *CASP8* | Caspase 8 | Encodes CASP8, involved in NF-κB activation or, alternatively, apoptosis. |
| *NFKB1* & *NFKB2* | Nuclear Factor Kappa B Subunit 1 and 2 | Encode NF-κB subunits, which contributes to the prevention of TNF-induced apoptosis through the expression of anti-apoptotic and inflammatory genes such as *TRAF1*, *TRAF2*, and *CASP8*. |

Supplementary Table 2. Summary-data-based Mendelian Randomization studies between TNF-related genes expression loci and Parkinson's disease risk and progression, and REM sleep behavior disorder risk

| Gene | topSNP | b.SMR | se.SMR | p.SMR | pFDR.SMR | p.multi SMR | pFDR.multi SMR | p.HEIDI | eQTL | Tissue | Disease |
|---|---|---|---|---|---|---|---|---|---|---|---|
| CASP8 | rs12990906 | 0.119664 | 0.131149 | 0.362 | 0.684 | 0.033 | 0.607 | 0.406 | BrainMeta | Brain Cortex | RBDrisk |
| NFKB1 | rs230541 | 0.145106 | 0.155188 | 0.35 | 0.684 | 0.35 | 0.752 | 0.495 | BrainMeta | Brain Cortex | RBDrisk |
| TNFRSF1A | rs4149577 | 0.0886967 | 0.252064 | 0.725 | 0.914 | 0.725 | 0.952 | 0.085 | BrainMeta | Brain Cortex | RBDrisk |
| TRAF1 | rs4310279 | 0.022075 | 0.0979041 | 0.822 | 0.914 | 0.976 | 0.976 | 0.931 | BrainMeta | Brain Cortex | RBDrisk |
| TRAF5 | rs2175633 | 0.187063 | 0.147401 | 0.204 | 0.623 | 0.204 | 0.701 | 0.351 | BrainMeta | Brain Cortex | RBDrisk |
| CASP8 | rs2540334 | -0.448773 | 0.403737 | 0.266 | 0.678 | 0.515 | 0.874 | 0.255 | BrainMeta | Brain Cortex | moca |
| NFKB1 | rs3774964 | 0.662164 | 0.408089 | 0.105 | 0.623 | 0.105 | 0.607 | 0.719 | BrainMeta | Brain Cortex | moca |
| TRAF1 | rs6478485 | -0.293944 | 0.312833 | 0.347 | 0.684 | 0.516 | 0.874 | 0.928 | BrainMeta | Brain Cortex | moca |
| TRAF5 | rs12095062 | -0.515413 | 0.646322 | 0.425 | 0.765 | 0.425 | 0.792 | 0.093 | BrainMeta | Brain Cortex | moca |
| CASP8 | rs2540334 | 0.185759 | 0.0840138 | 0.027 | 0.623 | 0.193 | 0.701 | 0.668 | BrainMeta | Brain Cortex | updrs3 |
| NFKB1 | rs3774964 | 0.0239799 | 0.0802935 | 0.765 | 0.914 | 0.765 | 0.952 | 0.405 | BrainMeta | Brain Cortex | updrs3 |
| NFKB2 | rs36226954 | 0.280081 | 0.358733 | 0.435 | 0.765 | 0.435 | 0.792 | 0.141 | BrainMeta | Brain Cortex | updrs3 |
| TRAF1 | rs6478485 | -0.00678228 | 0.0628061 | 0.914 | 0.914 | 0.531 | 0.874 | 0.422 | BrainMeta | Brain Cortex | updrs3 |
| TRAF5 | rs9887769 | 0.289495 | 0.149409 | 0.053 | 0.623 | 0.053 | 0.607 | 0.146 | BrainMeta | Brain Cortex | updrs3 |
| CASP8 | rs12990906 | 0.163257 | 0.145745 | 0.263 | 0.678 | 0.686 | 0.946 | 0.764 | BrainMeta | Brain Cortex | motorPD |
| NFKB1 | rs230541 | 0.213339 | 0.17085 | 0.212 | 0.623 | 0.212 | 0.701 | 0.338 | BrainMeta | Brain Cortex | motorPD |
| TNFRSF1A | rs4149577 | 0.134531 | 0.278831 | 0.629 | 0.871 | 0.629 | 0.917 | NA | BrainMeta | Brain Cortex | motorPD |
| TRAF1 | rs4310279 | -0.128322 | 0.103322 | 0.214 | 0.623 | 0.629 | 0.917 | 0.363 | BrainMeta | Brain Cortex | motorPD |
| TRAF5 | rs9887769 | 0.148638 | 0.160486 | 0.354 | 0.684 | 0.354 | 0.752 | 0.612 | BrainMeta | Brain Cortex | motorPD |
| CASP8 | rs2110690 | 0.109681 | 0.0828049 | 0.185 | 0.623 | 0.139 | 0.644 | 0.466 | GTEx | Brain Cortex | RBDrisk |
| CASP8 | rs2110690 | -0.274977 | 0.204904 | 0.18 | 0.623 | 0.002 | 0.102 | 0.236 | GTEx | Whole Blood | RBDrisk |
| TRAF1 | rs4310279 | 0.0347127 | 0.154016 | 0.822 | 0.914 | 0.822 | 0.952 | 0.793 | GTEx | Brain Caudate basal ganglia | RBDrisk |
| TRAF1 | rs4310279 | 0.0338337 | 0.15011 | 0.822 | 0.914 | 0.95 | 0.969 | 0.971 | GTEx | Brain Cortex | RBDrisk |
| TRAF1 | rs6478485 | -0.0914507 | 0.417746 | 0.827 | 0.914 | 0.599 | 0.917 | 0.593 | GTEx | Whole Blood | RBDrisk |
| TRAF2 | rs7854924 | 0.729355 | 0.468016 | 0.119 | 0.623 | 0.119 | 0.607 | 0.428 | GTEx | Whole Blood | RBDrisk |
| CASP8 | rs2110690 | 0.120428 | 0.203642 | 0.554 | 0.853 | 0.821 | 0.952 | 0.379 | GTEx | Brain Cortex | moca |
| CASP8 | rs2110690 | -0.30192 | 0.50923 | 0.553 | 0.853 | 0.647 | 0.917 | 0.821 | GTEx | Whole Blood | moca |
| TRAF1 | rs6478485 | -0.428312 | 0.45943 | 0.351 | 0.684 | 0.351 | 0.752 | NA | GTEx | Brain Cortex | moca |
| TRAF1 | rs6478485 | -0.999101 | 1.07165 | 0.351 | 0.684 | 0.351 | 0.752 | NA | GTEx | Whole Blood | moca |
| CASP8 | rs2110690 | 0.0780834 | 0.0424785 | 0.066 | 0.623 | 0.094 | 0.607 | 0.937 | GTEx | Brain Cortex | updrs3 |
| CASP8 | rs2110690 | -0.19576 | 0.10382 | 0.059 | 0.623 | 0.284 | 0.752 | 0.93 | GTEx | Whole Blood | updrs3 |

| | | | | | | | | | | | |
|---|---|---|---|---|---|---|---|---|---|---|---|
| TRAF1 | rs6478485 | -0.00988261 | 0.0915258 | 0.914 | 0.914 | 0.914 | 0.952 | NA | GTEx | Brain Cortex | updrs3 |
| TRAF1 | rs6478485 | -0.0230526 | 0.213497 | 0.914 | 0.914 | 0.914 | 0.952 | NA | GTEx | Whole Blood | updrs3 |
| CASP8 | rs2110690 | 0.0443771 | 0.0926974 | 0.632 | 0.871 | 0.915 | 0.952 | 0.983 | GTEx | Brain Cortex | motorPD |
| CASP8 | rs2110690 | -0.111256 | 0.232006 | 0.632 | 0.871 | 0.806 | 0.952 | 0.826 | GTEx | Whole Blood | motorPD |
| TRAF1 | rs4310279 | -0.201785 | 0.164471 | 0.22 | 0.623 | 0.22 | 0.701 | 0.189 | GTEx | Brain Caudate basal ganglia | motorPD |
| TRAF1 | rs4310279 | -0.196676 | 0.160129 | 0.219 | 0.623 | 0.392 | 0.792 | 0.737 | GTEx | Brain Cortex | motorPD |
| TRAF1 | rs6478485 | -0.263261 | 0.449534 | 0.558 | 0.853 | 0.825 | 0.952 | 0.575 | GTEx | Whole Blood | motorPD |
| TRAF2 | rs7854924 | 0.500918 | 0.478137 | 0.295 | 0.684 | 0.295 | 0.752 | NA | GTEx | Whole Blood | motorPD |
| CASP8 | rs12990906 | 0.0245294 | 0.0187634 | 0.191 | 0.623 | 0.565 | 0.9 | 0.679 | BrainMeta | Brain Cortex | PDrisk |
| NFKB1 | rs230541 | -0.0372947 | 0.0222532 | 0.094 | 0.623 | 0.094 | 0.607 | 0.353 | BrainMeta | Brain Cortex | PDrisk |
| NFKB2 | rs36226954 | 0.0507343 | 0.0317294 | 0.11 | 0.623 | 0.11 | 0.607 | 0.121 | BrainMeta | Brain Cortex | PDrisk |
| TNFRSF1A | rs4149577 | 0.0127618 | 0.0376632 | 0.735 | 0.914 | 0.735 | 0.952 | 0.756 | BrainMeta | Brain Cortex | PDrisk |
| TRAF1 | rs4310279 | 0.00206971 | 0.0129281 | 0.873 | 0.914 | 0.071 | 0.607 | 0.364 | BrainMeta | Brain Cortex | PDrisk |
| TRAF5 | rs2175633 | 0.040204 | 0.0203418 | 0.048 | 0.623 | 0.048 | 0.607 | 0.419 | BrainMeta | Brain Cortex | PDrisk |
| CASP8 | rs2110690 | 0.00643638 | 0.0113066 | 0.569 | 0.853 | 0.353 | 0.752 | 0.398 | GTEx | Brain Cortex | PDrisk |
| CASP8 | rs2110690 | -0.0161364 | 0.0282788 | 0.568 | 0.853 | 0.185 | 0.701 | 0.152 | GTEx | Whole Blood | PDrisk |
| TRAF1 | rs4310279 | 0.0032546 | 0.0203335 | 0.873 | 0.914 | 0.873 | 0.952 | 0.781 | GTEx | Brain Caudate basal ganglia | PDrisk |
| TRAF1 | rs4310279 | 0.00317219 | 0.0198182 | 0.873 | 0.914 | 0.421 | 0.792 | 0.554 | GTEx | Brain Cortex | PDrisk |
| TRAF1 | rs6478485 | -0.0774568 | 0.0596796 | 0.194 | 0.623 | 0.267 | 0.752 | 0.905 | GTEx | Whole Blood | PDrisk |
| TRAF2 | rs7854924 | -0.0089741 | 0.061741 | 0.884 | 0.914 | 0.884 | 0.952 | 0.626 | GTEx | Whole Blood | PDrisk |

SNP – Single Nucleotide Polymorphism; b – effect size (regression coefficient); SMR – Summary-data-based Mendelian Randomization; SE – Standard error; FDR – false discovery rate; eQTL – expression quantitative trait loci.

Supplementary Table 3. Rare variant association analysis in TNF-related genes in Parkinson’s disease cases and controls

| SetID | p.value | p.FDR | N.Marker |
|---|---|---|---|
| TNFRSF1B_ALL_UKBPD | 0.104 | 0.598 | 3329 |
| TRAF5_ALL_UKBPD | 0.447 | 0.855 | 3475 |
| CASP8_ALL_UKBPD | 1 | 1 | 1930 |
| NFKB1_ALL_UKBPD | 0.21 | 0.823 | 9006 |
| TNF_ALL_UKBPD | 0.757 | 0.873 | 186 |
| TRAF1_ALL_UKBPD | 0.746 | 0.873 | 1873 |
| TRAF2_ALL_UKBPD | 0.644 | 0.855 | 4444 |
| NFKB2_ALL_UKBPD | 0.672 | 0.866 | 511 |
| TNFRSF1A_ALL_UKBPD | 0.719 | 0.873 | 1036 |
| TRADD_ALL_UKBPD | 0.349 | 0.855 | 420 |
| TNFRSF1B_CADD_UKBPD | 0.613 | 0.855 | 8 |
| TRAF5_CADD_UKBPD | 0.031 | 0.598 | 42 |
| CASP8_CADD_UKBPD | 0.103 | 0.598 | 8 |
| NFKB1_CADD_UKBPD | 0.219 | 0.826 | 50 |
| TNF_CADD_UKBPD | 0.836 | 0.902 | 6 |
| TRAF1_CADD_UKBPD | 0.74 | 0.873 | 24 |
| TRAF2_CADD_UKBPD | 0.501 | 0.855 | 37 |
| NFKB2_CADD_UKBPD | 0.625 | 0.855 | 31 |
| TNFRSF1A_CADD_UKBPD | 1 | 1 | 9 |
| TRADD_CADD_UKBPD | 0.853 | 0.902 | 3 |
| TNFRSF1B_LOF_UKBPD | 0.62 | 0.855 | 3 |
| TRAF5_LOF_UKBPD | 0.681 | 0.866 | 14 |
| CASP8_LOF_UKBPD | 0.302 | 0.855 | 6 |
| NFKB1_LOF_UKBPD | 0.507 | 0.855 | 2 |
| TNF_LOF_UKBPD | 0.502 | 0.855 | 1 |
| TRAF1_LOF_UKBPD | 0.707 | 0.873 | 3 |
| TRAF2_LOF_UKBPD | 0.094 | 0.598 | 4 |
| NFKB2_LOF_UKBPD | 0.39 | 0.855 | 1 |
| TNFRSF1A_LOF_UKBPD | 0.537 | 0.855 | 2 |
| TNFRSF1B_MISSENSE_UKBPD | 0.559 | 0.855 | 71 |
| TRAF5_MISSENSE_UKBPD | 0.075 | 0.598 | 75 |
| CASP8_MISSENSE_UKBPD | 0.248 | 0.855 | 30 |
| NFKB1_MISSENSE_UKBPD | 0.2 | 0.823 | 148 |
| TNF_MISSENSE_UKBPD | 0.631 | 0.855 | 23 |
| TRAF1_MISSENSE_UKBPD | 0.561 | 0.855 | 69 |
| TRAF2_MISSENSE_UKBPD | 0.514 | 0.855 | 87 |
| NFKB2_MISSENSE_UKBPD | 0.588 | 0.855 | 13 |
| TNFRSF1A_MISSENSE_UKBPD | 0.421 | 0.855 | 50 |
| TRADD_MISSENSE_UKBPD | 0.362 | 0.855 | 43 |
| TNFRSF1B_ALL_AMPPD | 0.154 | 0.756 | 236 |

| | | | |
|---|---|---|---|
| TRAF5_ALL_AMPPD | 0.01 | 0.327 | 169 |
| CASP8_ALL_AMPPD | 0.307 | 0.855 | 86 |
| NFKB1_ALL_AMPPD | 0.293 | 0.855 | 474 |
| TNF_ALL_AMPPD | 0.181 | 0.808 | 17 |
| TRAF1_ALL_AMPPD | 0.557 | 0.855 | 137 |
| TRAF2_ALL_AMPPD | 0.376 | 0.855 | 252 |
| NFKB2_ALL_AMPPD | 0.282 | 0.855 | 50 |
| TNFRSF1A_ALL_AMPPD | 0.393 | 0.855 | 106 |
| TRADD_ALL_AMPPD | 0.773 | 0.875 | 27 |
| TRAF5_LOF_AMPPD | 0.617 | 0.855 | 1 |
| TNFRSF1B_MISSENSE_AMPPD | 0.097 | 0.598 | 12 |
| TRAF5_MISSENSE_AMPPD | 0.353 | 0.855 | 13 |
| CASP8_MISSENSE_AMPPD | 0.542 | 0.855 | 4 |
| NFKB1_MISSENSE_AMPPD | 0.73 | 0.873 | 18 |
| TNF_MISSENSE_AMPPD | 0.052 | 0.598 | 4 |
| TRAF1_MISSENSE_AMPPD | 0.006 | 0.312 | 17 |
| TRAF2_MISSENSE_AMPPD | 0.697 | 0.873 | 10 |
| NFKB2_MISSENSE_AMPPD | 0.878 | 0.905 | 2 |
| TNFRSF1A_MISSENSE_AMPPD | 0.053 | 0.598 | 12 |
| TRADD_MISSENSE_AMPPD | 0.499 | 0.855 | 2 |
| TNFRSF1B_CADD_AMPPD | 0.203 | 0.823 | 2 |
| TRAF5_CADD_AMPPD | 0.551 | 0.855 | 6 |
| CASP8_CADD_AMPPD | 0.777 | 0.875 | 2 |
| NFKB1_CADD_AMPPD | 0.102 | 0.598 | 9 |
| TNF_CADD_AMPPD | 0.281 | 0.855 | 2 |
| TRAF1_CADD_AMPPD | 0.057 | 0.598 | 10 |
| TRAF2_CADD_AMPPD | 0.578 | 0.855 | 6 |
| NFKB2_CADD_AMPPD | 0.275 | 0.855 | 8 |
| TNFRSF1A_CADD_AMPPD | 0.07 | 0.598 | 5 |
| CASP8_ALL_META | 1 | 1 | 2016 |
| CASP8_CADD_META | 0.114 | 0.621 | 10 |
| CASP8_MISSENSE_META | 0.37 | 0.855 | 34 |
| NFKB1_CADD_META | 0.645 | 0.855 | 59 |
| NFKB1_MISSENSE_META | 0.31 | 0.855 | 166 |
| NFKB2_ALL_META | 0.811 | 0.893 | 561 |
| NFKB2_CADD_META | 0.757 | 0.873 | 39 |
| NFKB2_MISSENSE_META | 0.865 | 0.902 | 15 |
| TNF_ALL_META | 0.665 | 0.866 | 203 |
| TNF_CADD_META | 0.864 | 0.902 | 8 |
| TNF_MISSENSE_META | 0.079 | 0.598 | 27 |
| TNFRSF1A_ALL_META | 0.371 | 0.855 | 1142 |
| TNFRSF1A_CADD_META | 0.513 | 0.855 | 14 |
| TNFRSF1A_MISSENSE_META | 0.523 | 0.855 | 62 |

| | | | |
|---|---|---|---|
| TNFRSF1B_ALL_META | 0.067 | 0.598 | 3565 |
| TNFRSF1B_CADD_META | 0.506 | 0.855 | 10 |
| TNFRSF1B_MISSENSE_META | 0.395 | 0.855 | 83 |
| TRADD_ALL_META | 0.633 | 0.855 | 447 |
| TRADD_MISSENSE_META | 0.5 | 0.855 | 45 |
| TRAF1_ALL_META | 0.854 | 0.902 | 2010 |
| TRAF1_CADD_META | 0.181 | 0.808 | 34 |
| TRAF1_MISSENSE_META | 0.228 | 0.826 | 86 |
| TRAF2_ALL_META | 0.533 | 0.855 | 4696 |
| TRAF2_CADD_META | 0.381 | 0.855 | 43 |
| TRAF2_MISSENSE_META | 0.404 | 0.855 | 97 |
| TRAF5_ALL_META | 0.128 | 0.662 | 3644 |
| TRAF5_CADD_META | 0.005 | 0.312 | 48 |
| TRAF5_LOF_META | 0.79 | 0.88 | 15 |
| TRAF5_MISSENSE_META | 0.037 | 0.598 | 88 |
| PATHWAY_CADD_UKBPD | 0.693 | 0.721 | 218 |
| PATHWAY_LOF_UKBPD | 0.484 | 0.721 | 36 |
| PATHWAY_MISSENSE_UKBPD | 0.149 | 0.346 | 609 |
| PATHWAY_CADD_AMPPD | 0.068 | 0.346 | 50 |
| PATHWAY_LOF_AMPPD | 0.617 | 0.721 | 1 |
| PATHWAY_MISSENSE_AMPPD | 0.154 | 0.346 | 94 |
| PATHWAY_CADD_META | 0.721 | 0.721 | 268 |
| PATHWAY_LOF_META | 0.661 | 0.721 | 37 |
| PATHWAY_MISSENSE_META | 0.113 | 0.346 | 703 |

FDR – false discovery rate; UKBPD – UK Biobank including Parkinson's disease cases and controls; AMPPD – Accelerating Medicines Partnership Parkinson's Disease; CADD – Combined Annotation Dependent Depletion (variants with score >20); LOF – loss-of-function; META – meta-analysis of the cohorts.

Supplementary Table 4. Rare variant association analysis in TNF-related genes in Parkinson's disease cases, proxy cases, and controls

| SetID | p.value | p.FDR | N.Marker |
|---|---|---|---|
| TNFRSF1B_ALL_AMPPD | 0.154 | 0.71 | 236 |
| TRAF5_ALL_AMPPD | 0.01 | 0.245 | 169 |
| CASP8_ALL_AMPPD | 0.307 | 0.753 | 86 |
| NFKB1_ALL_AMPPD | 0.293 | 0.753 | 474 |
| TNF_ALL_AMPPD | 0.181 | 0.71 | 17 |
| TRAF1_ALL_AMPPD | 0.557 | 0.805 | 137 |
| TRAF2_ALL_AMPPD | 0.376 | 0.77 | 252 |
| NFKB2_ALL_AMPPD | 0.282 | 0.753 | 50 |
| TNFRSF1A_ALL_AMPPD | 0.393 | 0.77 | 106 |
| TRADD_ALL_AMPPD | 0.773 | 0.856 | 27 |
| TRAF5_LOF_AMPPD | 0.617 | 0.805 | 1 |
| TNFRSF1B_MISSENSE_AMPPD | 0.097 | 0.555 | 12 |
| TRAF5_MISSENSE_AMPPD | 0.353 | 0.77 | 13 |
| CASP8_MISSENSE_AMPPD | 0.542 | 0.805 | 4 |
| NFKB1_MISSENSE_AMPPD | 0.73 | 0.822 | 18 |
| TNF_MISSENSE_AMPPD | 0.052 | 0.418 | 4 |
| TRAF1_MISSENSE_AMPPD | 0.006 | 0.228 | 17 |
| TRAF2_MISSENSE_AMPPD | 0.697 | 0.805 | 10 |
| NFKB2_MISSENSE_AMPPD | 0.878 | 0.935 | 2 |
| TNFRSF1A_MISSENSE_AMPPD | 0.053 | 0.418 | 12 |
| TRADD_MISSENSE_AMPPD | 0.499 | 0.805 | 2 |
| TNFRSF1B_CADD_AMPPD | 0.203 | 0.753 | 2 |
| TRAF5_CADD_AMPPD | 0.551 | 0.805 | 6 |
| CASP8_CADD_AMPPD | 0.777 | 0.856 | 2 |
| NFKB1_CADD_AMPPD | 0.102 | 0.555 | 9 |
| TNF_CADD_AMPPD | 0.281 | 0.753 | 2 |
| TRAF1_CADD_AMPPD | 0.057 | 0.418 | 10 |
| TRAF2_CADD_AMPPD | 0.578 | 0.805 | 6 |
| NFKB2_CADD_AMPPD | 0.275 | 0.753 | 8 |
| TNFRSF1A_CADD_AMPPD | 0.07 | 0.456 | 5 |
| CASP8_ALL_META | 0.007 | 0.228 | 2214 |
| TNF_ALL_META | 0.544 | 0.805 | 221 |
| TNFRSF1B_ALL_META | 0.811 | 0.883 | 3899 |
| TRAF5_ALL_META | 0.18 | 0.71 | 3990 |
| CASP8_CADD_META | 0.652 | 0.805 | 12 |
| CASP8_MISSENSE_META | 0.715 | 0.815 | 38 |
| NFKB1_CADD_META | 0.598 | 0.805 | 62 |
| NFKB1_MISSENSE_META | 0.457 | 0.804 | 180 |
| NFKB2_ALL_META | 0.295 | 0.753 | 605 |
| NFKB2_CADD_META | 0.167 | 0.71 | 41 |

| | | | |
|---|---|---|---|
| NFKB2_MISSENSE_META | 0.296 | 0.753 | 19 |
| TNF_CADD_META | 1 | 1 | 10 |
| TNF_MISSENSE_META | 0.347 | 0.77 | 35 |
| TNFRSF1A_ALL_META | 0.887 | 0.935 | 1262 |
| TNFRSF1A_CADD_META | 0.35 | 0.77 | 14 |
| TNFRSF1A_MISSENSE_META | 0.665 | 0.805 | 68 |
| TNFRSF1B_CADD_META | 0.672 | 0.805 | 12 |
| TNFRSF1B_MISSENSE_META | 0.852 | 0.918 | 93 |
| TRADD_ALL_META | 0.372 | 0.77 | 499 |
| TRADD_MISSENSE_META | 0.476 | 0.804 | 47 |
| TRAF1_ALL_META | 0.29 | 0.753 | 2204 |
| TRAF1_CADD_META | 0.175 | 0.71 | 35 |
| TRAF1_MISSENSE_META | 0.252 | 0.753 | 90 |
| TRAF2_ALL_META | 0.433 | 0.785 | 5111 |
| TRAF2_CADD_META | 0.174 | 0.71 | 47 |
| TRAF2_MISSENSE_META | 0.034 | 0.418 | 105 |
| TRAF5_CADD_META | 0.032 | 0.418 | 51 |
| TRAF5_LOF_META | 0.356 | 0.77 | 17 |
| TRAF5_MISSENSE_META | 0.469 | 0.804 | 97 |
| TNFRSF1B_ALL_UKBPPROXY | 0.937 | 0.977 | 3663 |
| TRAF5_ALL_UKBPPROXY | 0.564 | 0.805 | 3821 |
| CASP8_ALL_UKBPPROXY | 0.007 | 0.228 | 2128 |
| TNF_ALL_UKBPPROXY | 0.504 | 0.805 | 204 |
| TRAF1_ALL_UKBPPROXY | 0.06 | 0.418 | 2067 |
| TRAF2_ALL_UKBPPROXY | 0.272 | 0.753 | 4859 |
| NFKB2_ALL_UKBPPROXY | 0.255 | 0.753 | 555 |
| TNFRSF1A_ALL_UKBPPROXY | 1 | 1 | 1156 |
| TRADD_ALL_UKBPPROXY | 0.345 | 0.77 | 472 |
| TNFRSF1B_CADD_UKBPPROXY | 0.459 | 0.804 | 10 |
| TRAF5_CADD_UKBPPROXY | 0.045 | 0.418 | 45 |
| CASP8_CADD_UKBPPROXY | 0.65 | 0.805 | 10 |
| NFKB1_CADD_UKBPPROXY | 0.413 | 0.779 | 53 |
| TNF_CADD_UKBPPROXY | 1 | 1 | 8 |
| TRAF1_CADD_UKBPPROXY | 0.385 | 0.77 | 25 |
| TRAF2_CADD_UKBPPROXY | 0.232 | 0.753 | 41 |
| NFKB2_CADD_UKBPPROXY | 0.086 | 0.527 | 33 |
| TNFRSF1A_CADD_UKBPPROXY | 0.66 | 0.805 | 9 |
| TRADD_CADD_UKBPPROXY | 0.386 | 0.77 | 3 |
| TNFRSF1B_LOF_UKBPPROXY | 0.653 | 0.805 | 4 |
| TRAF5_LOF_UKBPPROXY | 0.305 | 0.753 | 16 |
| CASP8_LOF_UKBPPROXY | 0.639 | 0.805 | 8 |
| NFKB1_LOF_UKBPPROXY | 0.057 | 0.418 | 4 |
| TNF_LOF_UKBPPROXY | 0.567 | 0.805 | 1 |

| | | | |
|---|---|---|---|
| TRAF1_LOF_UKBPPROXY | 0.401 | 0.771 | 3 |
| TRAF2_LOF_UKBPPROXY | 0.026 | 0.418 | 7 |
| NFKB2_LOF_UKBPPROXY | 1 | 1 | 1 |
| TNFRSF1A_LOF_UKBPPROXY | 0.647 | 0.805 | 3 |
| TRADD_LOF_UKBPPROXY | 0.136 | 0.704 | 1 |
| TNFRSF1B_MISSENSE_UKBPPROXY | 0.601 | 0.805 | 81 |
| TRAF5_MISSENSE_UKBPPROXY | 0.555 | 0.805 | 84 |
| CASP8_MISSENSE_UKBPPROXY | 0.627 | 0.805 | 34 |
| NFKB1_MISSENSE_UKBPPROXY | 0.424 | 0.784 | 162 |
| TNF_MISSENSE_UKBPPROXY | 0.698 | 0.805 | 31 |
| TRAF1_MISSENSE_UKBPPROXY | 0.677 | 0.805 | 73 |
| TRAF2_MISSENSE_UKBPPROXY | 0.045 | 0.418 | 95 |
| NFKB2_MISSENSE_UKBPPROXY | 0.224 | 0.753 | 17 |
| TNFRSF1A_MISSENSE_UKBPPROXY | 0.682 | 0.805 | 56 |
| TRADD_MISSENSE_UKBPPROXY | 0.566 | 0.805 | 45 |
| PATHWAY_MISSENSE_UKBPPROXY | 0.791 | 0.791 | 678 |
| PATHWAY_LOF_UKBPPROXY | 0.198 | 0.338 | 48 |
| PATHWAY_CADD_UKBPPROXY | 0.117 | 0.338 | 237 |
| PATHWAY_CADD_AMPPD | 0.068 | 0.338 | 50 |
| PATHWAY_LOF_AMPPD | 0.617 | 0.791 | 1 |
| PATHWAY_MISSENSE_AMPPD | 0.154 | 0.338 | 94 |
| PATHWAY_CADD_META | 0.159 | 0.338 | 287 |
| PATHWAY_LOF_META | 0.225 | 0.338 | 49 |
| PATHWAY_MISSENSE_META | 0.723 | 0.791 | 772 |

FDR – false discovery rate; UKBPROXY – UK Biobank including Parkinson's disease cases, proxy cases, and controls; AMPPD – Accelerating Medicines Partnership Parkinson's Disease; CADD – Combined Annotation Dependent Depletion (variants with score >20); LOF – loss-of-function; META – meta-analysis of the cohorts.

Supplementary Table 5. Study population for pathway specific polygenic risk score analysis of TNF-related genes

| Cohort | N_controls | N_cases | N_Male | N_Female | Mean_age |
|---|---|---|---|---|---|
| McGill | 2127 | 3242 | 3098 | 2271 | 55.88 |
| PPMI | 164 | 417 | 387 | 194 | 60.18 |
| APDGC | 302 | 621 | 599 | 324 | 78.89 |
| IPDGC | 5480 | 5229 | 6421 | 4288 | 62.78 |
| NINDS | 790 | 896 | 865 | 821 | 62.64 |
| NGRC | 1968 | 1972 | 2092 | 1848 | 64.29 |
| UKB | 65720 | 3286 | 33999 | 35007 | 63.84 |

PPMI- Parkinson's Progression Markers Initiative; APDGC- Autopsy-Confirmed Parkinson Disease GWAS Consortium; IPDGC -International Parkinson Disease Genomics Consortium; NINDS- National Institute of Neurological Disorders and Stroke Repository Parkinson's Disease Collection; NGRC- NeuroGenetics Research Consortium; UKB-UK Biobank

Supplementary Table 6. Pathway specific polygenic risk score analysis of TNF-related genes in Parkinson's disease

| Cohort | P | OR | SE | 95%CI |
|---|---|---|---|---|
| McGill | 0.000491 | 0.895308 | 0.028407352 | [0.8413; 0.9528] |
| PPMI | 0.500551 | 0.93519 | 0.093023952 | [0.7695; 1.1365] |
| APDGC | 0.957311 | 0.996073 | 0.07322568 | [0.8624; 1.1504] |
| IPDGC | 0.468675 | 1.014351 | 0.019945826 | [0.9760; 1.0542] |
| NINDS | 0.110751 | 1.086555 | 0.056556286 | [0.9812; 1.2033] |
| NGRC | 0.592286 | 0.980538 | 0.035986092 | [0.9125; 1.0537] |
| UKB | 0.046068 | 1.039793 | 0.020340316 | [1.0007; 1.0804] |
| Random effects model | 0.8404 | 0.9951 | | [0.9482; 1.0442] |

PPMI- Parkinson's Progression Markers Initiative; APDGC- Autopsy-Confirmed Parkinson Disease GWAS Consortium; IPDGC -International Parkinson Disease Genomics Consortium; NINDS- National Institute of Neurological Disorders and Stroke Repository Parkinson's Disease Collection; NGRC- NeuroGenetics Research Consortium; UKB-UK Biobank; OR- odds ratio; SE-standard error; CI- confidence interval
